# Wealth-Related Inequalities in Cesarean Section Utilization Among Facility-Based Births in Bangladesh: Evidence from Public and Private Healthcare Facilities

**DOI:** 10.64898/2026.06.09.26355308

**Authors:** Sultan Mahmud

**Affiliations:** Maternal and Child Health Division, International Centre for Diarrhoeal Disease Research, Dhaka 1212, Bangladesh

**Keywords:** Cesarean section, Socioeconomic inequality, Concentration index, Decomposition analysis, Public facilities, Private facilities, Bangladesh

## Abstract

**Background:** Bangladesh has experienced a rapid increase in cesarean section (CS) utilization over the past two decades. While previous studies have documented socioeconomic disparities in CS use, evidence on how wealth-related inequalities differ between public and private healthcare facilities remains limited. This study assessed the magnitude and drivers of socioeconomic inequality in CS utilization among facility-based births in Bangladesh.

**Methods:** We analyzed data from 3,008 facility-based births reported in the 2022 Bangladesh Demographic and Health Survey (BDHS). Survey-weighted multivariable logistic regression was used to identify factors associated with CS utilization. Wealth-related inequality was assessed using concentration curves and the Erreygers-corrected concentration index (ECCI). Regression-based decomposition of the standard concentration index was performed to quantify the contribution of socioeconomic, demographic, and healthcare-related factors to observed inequalities overall and separately for public and private facilities.

**Results:** Overall, 71.2% of facility-based births were delivered by CS, with substantially higher prevalence in private facilities (84.2%) than in public facilities (35.9%). Women delivering in private facilities had markedly higher odds of CS than those delivering in public facilities (adjusted odds ratio [AOR]: 9.07; 95% confidence interval [CI]: 7.17–11.47). Significant pro-rich inequality was observed overall (ECCI: 0.154; 95% CI: 0.117–0.191), with inequality substantially greater in public facilities (ECCI: 0.189; 95% CI: 0.114–0.264) than in private facilities (ECCI: 0.049; 95% CI: 0.014–0.084). Decomposition analysis showed that household wealth was the dominant contributor to inequality, particularly the richest wealth quintile, accounting for 81.5% of overall inequality, 63.8% in public facilities, and 109.7% in private facilities.

**Conclusions:** Wealth-related inequalities in CS utilization remain substantial in Bangladesh despite widespread use of the procedure. Although pro-rich inequality exists across both sectors, inequality is considerably greater in public facilities and is driven by different mechanisms across facility types. Policies should simultaneously improve equitable access to medically necessary CS and reduce unnecessary procedures, particularly within the private sector.

## Introduction

Cesarean section (CS) is a critical component of comprehensive obstetric care and can substantially reduce maternal and neonatal morbidity and mortality when medically indicated. Access to timely CS is particularly important for managing obstetric emergencies such as obstructed labor, fetal distress, placenta previa, and other life-threatening complications [1]. However, increasing evidence suggests that the global challenge is no longer limited to inadequate access [2]. Many countries now face a dual burden characterized by underuse of medically necessary CS among disadvantaged populations and overuse among socioeconomically advantaged groups [3, 4]. Both extremes have important consequences: insufficient access contributes to preventable maternal and neonatal deaths, whereas unnecessary procedures increase the risk of surgical complications, adverse reproductive outcomes, and avoidable healthcare expenditures [4, 5].

Over the past three decades, CS rates have risen dramatically worldwide, particularly in low– and middle-income countries (LMICs). Although part of this increase reflects improved access to emergency obstetric services, a growing body of evidence suggests that the expansion has been unevenly distributed across socioeconomic groups. Analyses from 72 LMICs demonstrated substantial within-country wealth-related inequalities in CS utilization, with the poorest women consistently exhibiting the lowest rates and the wealthiest women the highest rates, often indicating simultaneous underuse and overuse within the same population (<u>Boatin et al. 2018</u>). Similar findings have been reported across sub-Saharan Africa and South Asia, where household wealth, maternal education, and access to healthcare services remain powerful determinants of CS utilization [6, 7].

Bangladesh represents one of the most striking examples of this global phenomenon. Over the last two decades, the country has achieved remarkable improvements in maternal health service utilization, institutional delivery, and antenatal care coverage. During the same period, Bangladesh experienced a remarkable rise in CS utilization, with the proportion of births delivered by CS increasing from approximately 5% in 2004 to 45% in 2022 [8]. While this trend may indicate improved availability of obstetric care, concerns have emerged regarding the unequal distribution of CS services and the growing role of non-clinical factors in delivery decisions. Several studies have shown that women from wealthier households, those with higher educational attainment, and those delivering in private healthcare facilities are substantially more likely to undergo CS than their socioeconomically disadvantaged counterparts [5, 9, 10]. Consequently, the rapid rise in CS utilization in Bangladesh raises important questions regarding equity, appropriate use, and access to medically necessary obstetric interventions.

Recent evidence further indicates that inequalities in CS utilization remain persistent and, in some cases, have widened over time. Using WHO Health Equity Assessment Toolkit (HEAT) measures, Kundu et al. (2024) documented substantial socioeconomic and geographical disparities in CS use in Bangladesh [5], with utilization disproportionately concentrated among affluent women and urban populations. Similar patterns have been observed internationally, where wealth-related inequalities continue to characterize access to obstetric services despite overall increases in national CS rates [4, 6]. These findings suggest that national averages may conceal important inequities in service utilization and that rising CS rates do not necessarily translate into equitable access to essential obstetric care.

Although previous studies have identified several determinants of CS utilization, understanding the factors associated with CS use is not equivalent to understanding the drivers of socioeconomic inequality. Conventional regression analyses estimate associations between individual characteristics and CS utilization but provide limited insight into how these factors collectively generate wealth-related disparities [5, 11, 12]. Decomposition techniques offer an important advantage by quantifying the relative contribution of different socioeconomic, demographic, and healthcare-related factors to observed inequalities. Such evidence is particularly valuable for policy formulation because interventions targeting major contributors to inequality are more likely to reduce inequitable service utilization than those addressing individual risk factors alone.

Despite the growing body of literature on CS utilization in Bangladesh, evidence based on the most recent nationally representative data remains limited. Moreover, existing studies have primarily focused on identifying determinants of CS use or assessing overall socioeconomic inequalities, providing limited insight into the factors driving these disparities. Although a few studies have applied decomposition techniques to examine wealth-related inequalities in CS utilization, they have largely treated the population as a homogeneous group and have not explored whether the magnitude and underlying drivers of inequality differ between public and private healthcare facilities. Given the rapid expansion of the private health sector and its increasing role in childbirth care in Bangladesh, understanding facility-specific patterns of socioeconomic inequality is essential for designing targeted and equitable interventions.

Therefore, this study aimed to assess wealth-related socioeconomic inequalities in CS utilization among facility-based deliveries in Bangladesh using data from the 2022 Bangladesh Demographic and Health Survey. Specifically, we quantified the extent of inequality using concentration curves and the Erreygers corrected concentration index and decomposed the observed inequalities separately for public and private healthcare facilities to identify the relative contributions of socioeconomic, demographic, maternal, and healthcare-related factors. By uncovering the key drivers of inequality across facility types, this study provides evidence to inform policies aimed at promoting equitable access to medically necessary cesarean delivery while reducing unnecessary use of the procedure.

## Methods

### Study design and data source

This study utilized data from the 2022 Bangladesh Demographic and Health Survey (BDHS). The BDHS 2022 employed a stratified two-stage cluster sampling design based on the sampling frame derived from the 2011 Population and Housing Census conducted by the Bangladesh Bureau of Statistics (BBS). In the first stage, 675 enumeration areas (EAs)—including 237 urban and 438 rural EAs—were selected using probability proportional to size sampling. In the second stage, a systematic sample of 45 households from each selected EA was drawn. This sampling design was intended to provide nationally representative estimates of key demographic and health indicators for urban and rural populations separately and for each of the eight administrative divisions of Bangladesh.

All ever-married women aged 15–49 years residing in the selected households were eligible for individual interviews. Information was collected on a wide range of topics, including reproductive and maternal health, child health, healthcare utilization, fertility preferences, and socioeconomic and demographic characteristics. Further details regarding the survey methodology and sampling procedures are available in the BDHS 2022 final report [8].

### Study population

The analysis was restricted to women aged 15–49 years who had a live birth during the three years preceding the survey. A total of 4,818 women reported a recent live birth in the BDHS 2022 (Fig 1). Of these, 1,802 births that occurred outside health facilities were excluded because the study focused on facility-based deliveries. An additional eight observations with missing information on study variables were excluded through complete-case analysis. The final analytical sample comprised 3,008 facility-based births, including deliveries occurring in both public and private healthcare facilities.

**Fig 1.**
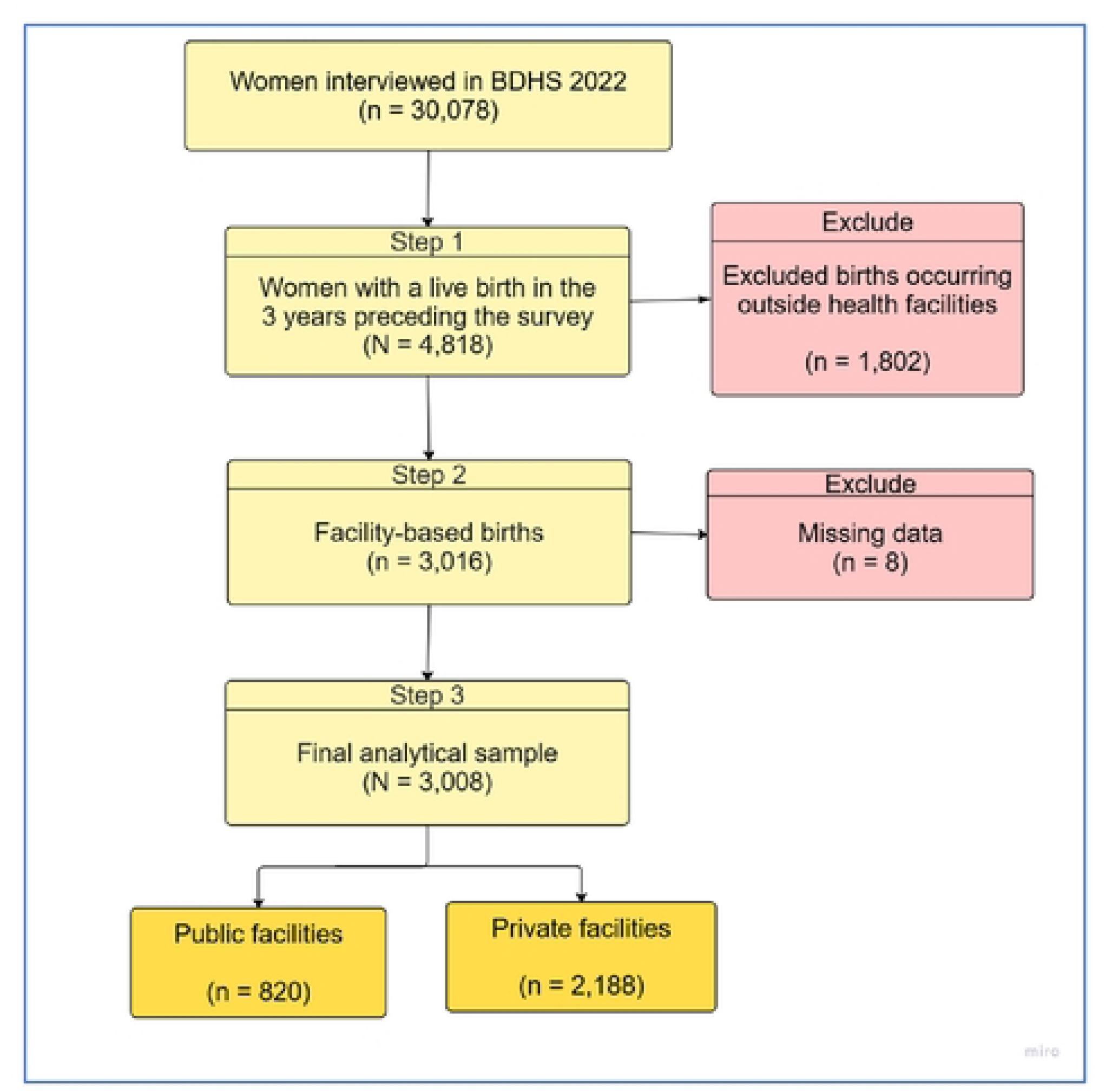
Flowchart of participant selection for the analysis of wealth-related inequalities in cesarean delivery among facility-based births, BDHS 2022.

### Outcome variable

The outcome variable was cesarean delivery, defined using the BDHS question asking whether the birth was delivered by CS. The women who delivered her last baby by cesarean section were coded as 1, while those reporting vaginal delivery were coded as 0.

### Explanatory variables

The primary explanatory variable was facility type, classified as either public or private according to the reported place of delivery. Public facilities included district hospitals, maternal and child welfare centres, upazila health complexes, union health and family welfare centres, community clinics, and other government health facilities. Private facilities included private hospitals, private clinics, nongovernmental organization (NGO) facilities, and other private-sector institutions. Facility type was included as an explanatory variable in the overall regression model and was also used to stratify the inequality and decomposition analyses.

Selection of explanatory variables was informed by previous literature on the determinants of cesarean delivery and the availability of relevant information in the BDHS 2022 dataset. The covariates included maternal age (15–20, 21–25, 26–30, and ≥31 years), maternal education (No education, Primary, Secondary, Higher), place of residence (urban and rural), religion (Islam and other religions), household wealth index (poorest, poorer, middle, richer, and richest), and administrative division (Barishal, Chattogram, Dhaka, Khulna, Mymensingh, Rajshahi, Rangpur, and Sylhet). Additional household and socioeconomic characteristics included husband’s education (<Secondary and ≥Secondary), husband’s occupation (not working, agricultural, manual labor, and Professional/Service/Business), media exposure (no and yes), and women’s employment status (no and yes).

Variables related to women’s autonomy and healthcare access included participation in household decision-making (no and yes) and whether distance to a health facility was reported as a problem in obtaining medical care (no and yes). Reproductive and maternal health characteristics comprised antenatal care (ANC) visits (<4 and ≥4 visits), parity (≤1, 2–3, and ≥4 children), and pregnancy intention (wanted then, wanted later, and wanted no more).

### Missing Data

Missing values were minimal (<1%) and were observed only for husband’s education (0.95%), husband’s occupation (0.92%), and participation in household decision-making (0.76%). Because the extent of missingness was negligible, records with missing information were excluded using complete-case analysis. The final analytical sample consisted of 3,008 facility-based births.

### Measurement of socioeconomic inequality

Socioeconomic inequality in cesarean delivery was assessed using concentration curves and the Erreygers-corrected concentration index (ECCI). Concentration curves plot the cumulative proportion of cesarean deliveries against the cumulative proportion of women ranked by household wealth from the poorest to the richest. If the concentration curve coincides with the 45° line of equality, cesarean delivery is equally distributed across wealth groups. A curve below the line of equality indicates that cesarean delivery is concentrated among wealthier women, whereas a curve above the line indicates concentration among poorer women.

The standard concentration index (CIx) was calculated as:

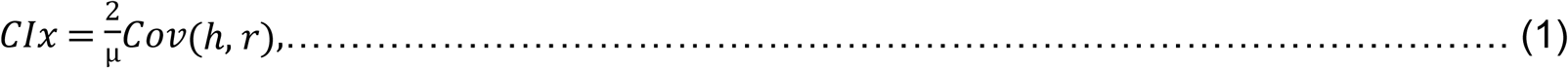

where ℎ denotes the health outcome (cesarean delivery), *r* is the fractional rank of individuals according to household wealth, µ is the mean of the outcome variable, and *Cov* represents the covariance between the outcome and wealth rank.

The concentration index ranges from −1 to +1. A positive value indicates a pro-rich distribution of cesarean delivery, whereas a negative value indicates a pro-poor distribution.

Because cesarean delivery is a binary outcome, the conventional concentration index is sensitive to the bounded nature of the variable. Therefore, the Erreygers correction was applied to obtain a normalized measure of inequality:

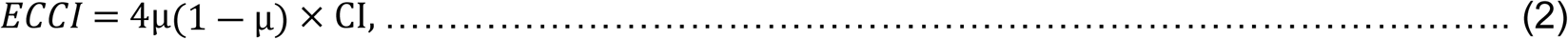

where *ECCI* is the Erreygers-corrected concentration index and µ is the mean prevalence of cesarean delivery. Positive values of *ECCI* indicate that cesarean delivery is disproportionately concentrated among wealthier women, whereas negative values indicate concentration among poorer women. Statistical significance was evaluated using 95% confidence intervals and *P-values*<0.05.

### Decomposition of socioeconomic inequality

To identify the determinants contributing to wealth-related inequality in cesarean delivery, the CIx was decomposed using the regression-based decomposition approach proposed by Wagstaff and subsequently adapted for the Erreygers index. The health outcome was expressed as a linear function of a set of explanatory variables:

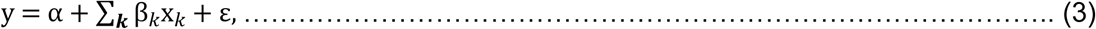

where *y* represents cesarean delivery status, *x* denotes the explanatory variables, *β* represents the regression coefficients, and *ε* is the error term. The contribution of *k^th^* determinant to the overall concentration index was calculated as:

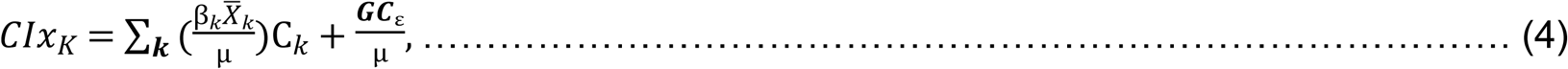

Where *CIx* is the concentration index of cesarean delivery, 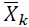 is the mean of determinant *k*, C_*k*_ is the concentration index of determinant *k*, and *GC*_ε_ is the generalized concentration index of the residual term. The elasticity of *k^th^* determinant was calculated as: 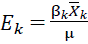, The absolute contribution of determinant *k: C*_*K*_ = *E*_*k*_ × *CIx*_*K*_ *and* the percentage contribution of *k^th^* determinant: 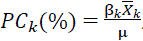. Where the positive percentage contributions indicate factors that increase pro-rich inequality in cesarean delivery, whereas negative contributions indicate factors that reduce the observed inequality.

### Statistical analysis

All statistical analyses were conducted using Stata/SE 16.0 (StataCorp LLC, College Station, TX, USA). Concentration curves, Erreygers-corrected concentration indices, and decomposition analyses were performed using R (R Foundation for Statistical Computing, Vienna, Austria). All analyses accounted for the complex survey design of the BDHS by incorporating sampling weights, clustering, and stratification.

Descriptive statistics were used to summarize participants’ characteristics overall and by facility type. Differences in the distribution of explanatory variables between public and private facility users were assessed using the Rao–Scott adjusted chi-square test. Multicollinearity among explanatory variables was evaluated using the variance inflation factor (VIF), and no evidence of problematic multicollinearity was observed (all VIFs < 10). Statistical significance was assessed at a two-sided p-value < 0.05. No variable selection procedure was performed because the study aimed to evaluate the contribution of all selected determinants to cesarean delivery and its socioeconomic inequality.

Survey-weighted multivariable logistic regression models were fitted to identify factors associated with cesarean delivery. Three separate models were estimated: (i) an overall model including all facility-based births, with facility type included as an explanatory variable; (ii) a model restricted to births occurring in public health facilities; and (iii) a model restricted to births occurring in private health facilities. Adjusted odds ratios (AORs) and 95% confidence intervals (CIs) were reported.

As cesarean delivery is a binary outcome, overall inequality was summarized using the Erreygers-corrected concentration index (ECCI), whereas decomposition was performed using the standard concentration index following the conventional Wagstaff decomposition approach.

This study was reported in accordance with the Strengthening the Reporting of Observational Studies in Epidemiology (STROBE) guidelines.

### Ethical considerations

The 2022 BDHS was approved by the Institutional Review Board of ICF International and the Bangladesh Medical Research Council (BMRC). Written informed consent was obtained from all participants prior to data collection.

This study was based on a secondary analysis of the BDHS dataset, which is publicly available upon request through the DHS Program. The dataset is fully anonymized and contains no personally identifiable information. Access to the data was granted by the DHS Program on 10 February 2026. The authors did not have access to any information that could identify individual participants at any stage of the study. Consequently, no additional ethical approval was required for this secondary data analysis. All analyses were conducted in accordance with the ethical principles governing the use of publicly available, de-identified survey data.

## Findings

A total of 3,008 facility-based births were included in the analysis. Based on weighted estimates, 26.9% of births occurred in public facilities and 73.1% in private facilities. Overall, the largest proportion of women were aged 21–25 years (34.1%), had secondary education (56.1%), resided in rural areas (69.7%), and were of the Islamic faith (90.7%). Approximately one-quarter belonged to the richest wealth quintile (25.2%), half had received four or more antenatal care (ANC) visits (50.3%), and 86.3% participated in household decision-making. In addition, 67.0% of women had husbands with secondary or higher education, and 54.7% had husbands engaged in professional, service, or business occupations.

Significant differences in socioeconomic and demographic characteristics were observed between women delivering in public and private facilities. Compared with women delivering in public facilities, those delivering in private facilities were more likely to have higher educational attainment (26.6% vs. 18.0% with higher education; p=0.002), belong to the richer or richest wealth quintiles (52.6% vs. 39.5%; p<0.001), have husbands with secondary or higher education (69.9% vs. 59.0%; p<0.001), and have husbands engaged in professional, service, or business occupations (57.8% vs. 46.4%; p<0.001). Women delivering in private facilities were also more likely to report media exposure (67.1% vs. 62.1%; p=0.046), participation in household decision-making (87.2% vs. 84.1%; p=0.048), and receipt of four or more ANC visits (52.3% vs. 44.8%; p=0.002). Furthermore, the distribution of births differed significantly across household wealth quintiles and administrative divisions (both p<0.001). Women delivering in private facilities also had lower parity (≥4 children: 4.4% vs. 7.0%; p=0.042). No significant differences were observed between public and private facility users with respect to maternal age, place of residence, religion, employment status, distance to health facilities, or pregnancy intention (all p>0.05).

**Table 1.**
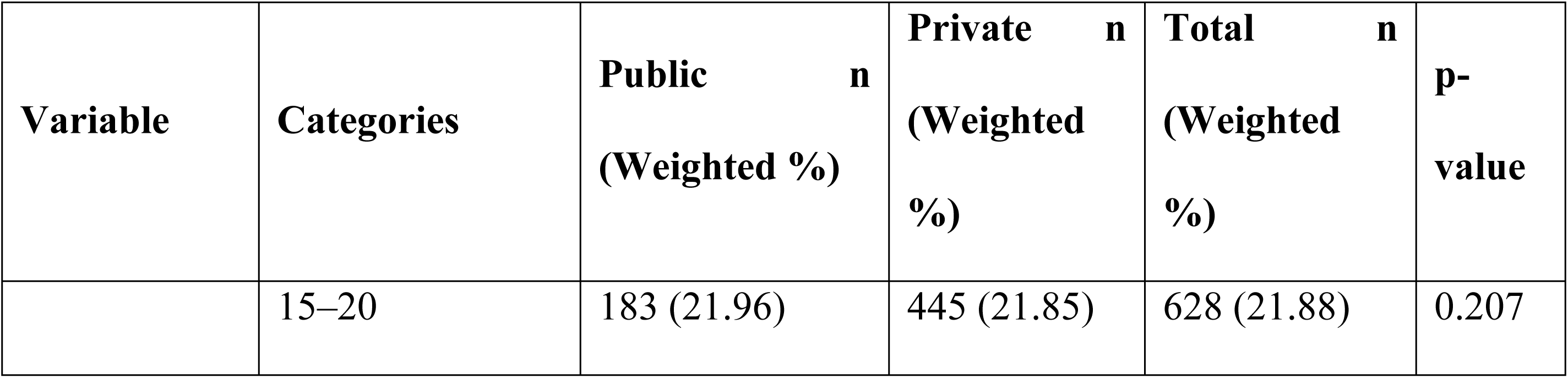
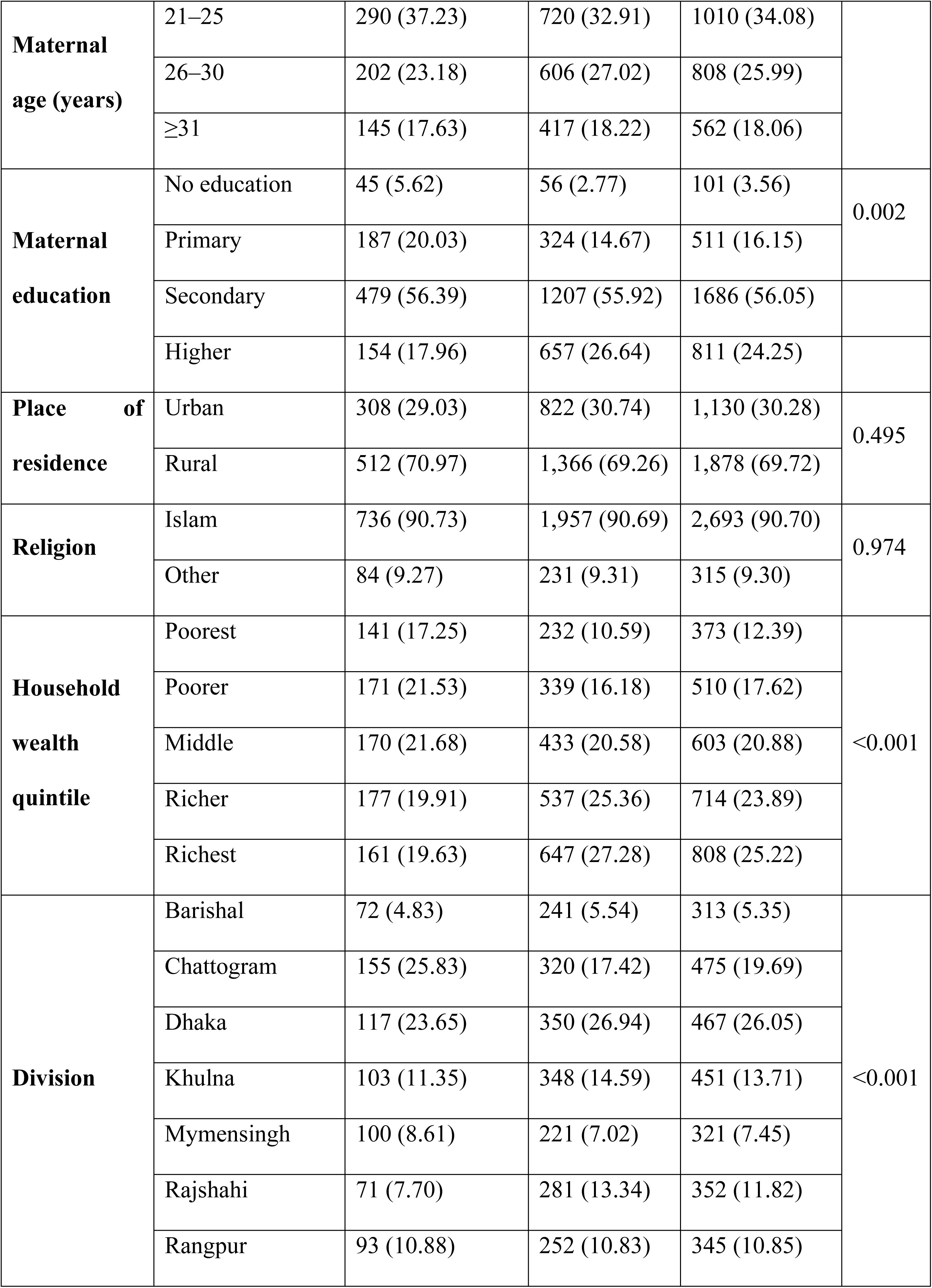
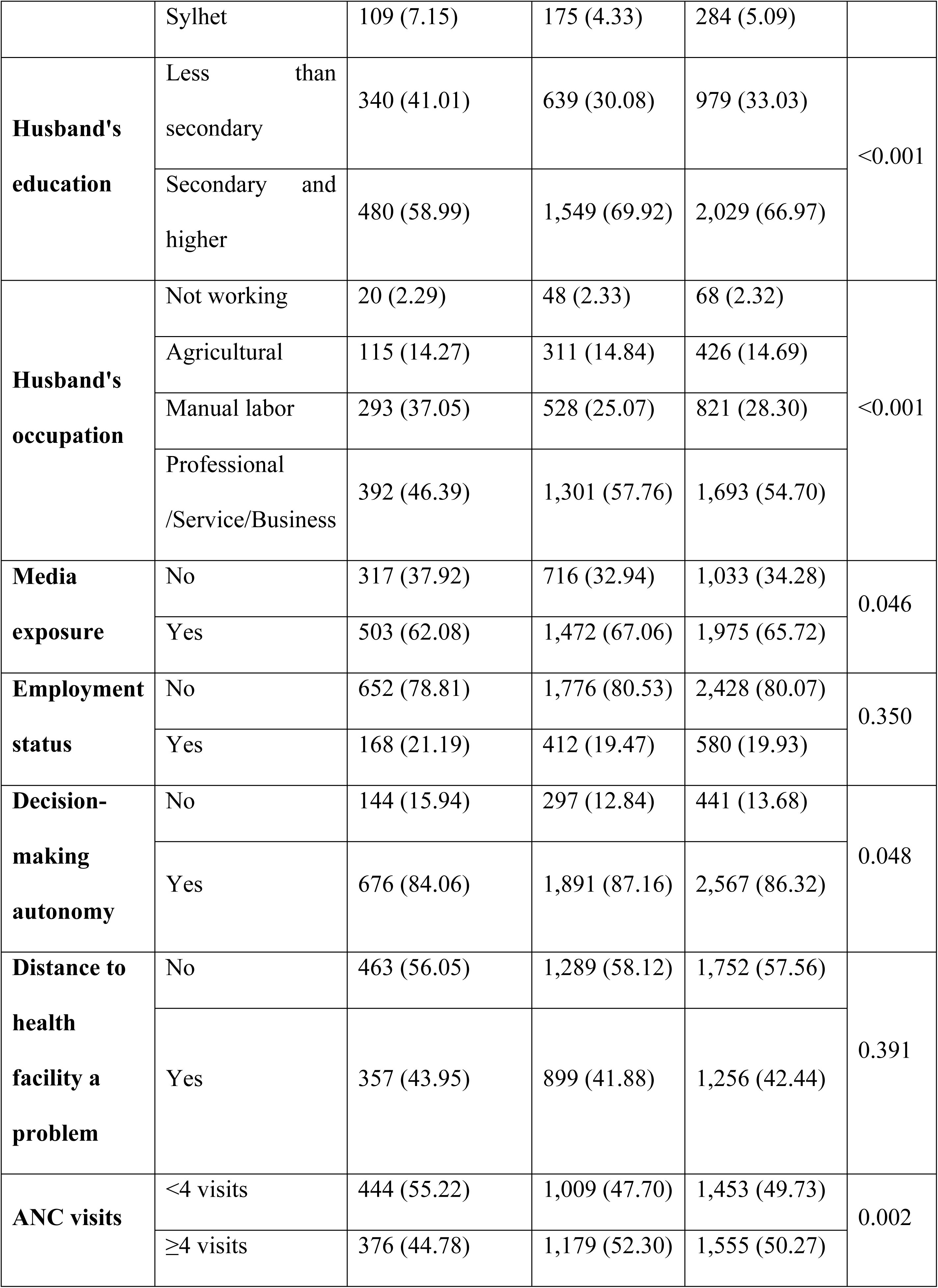
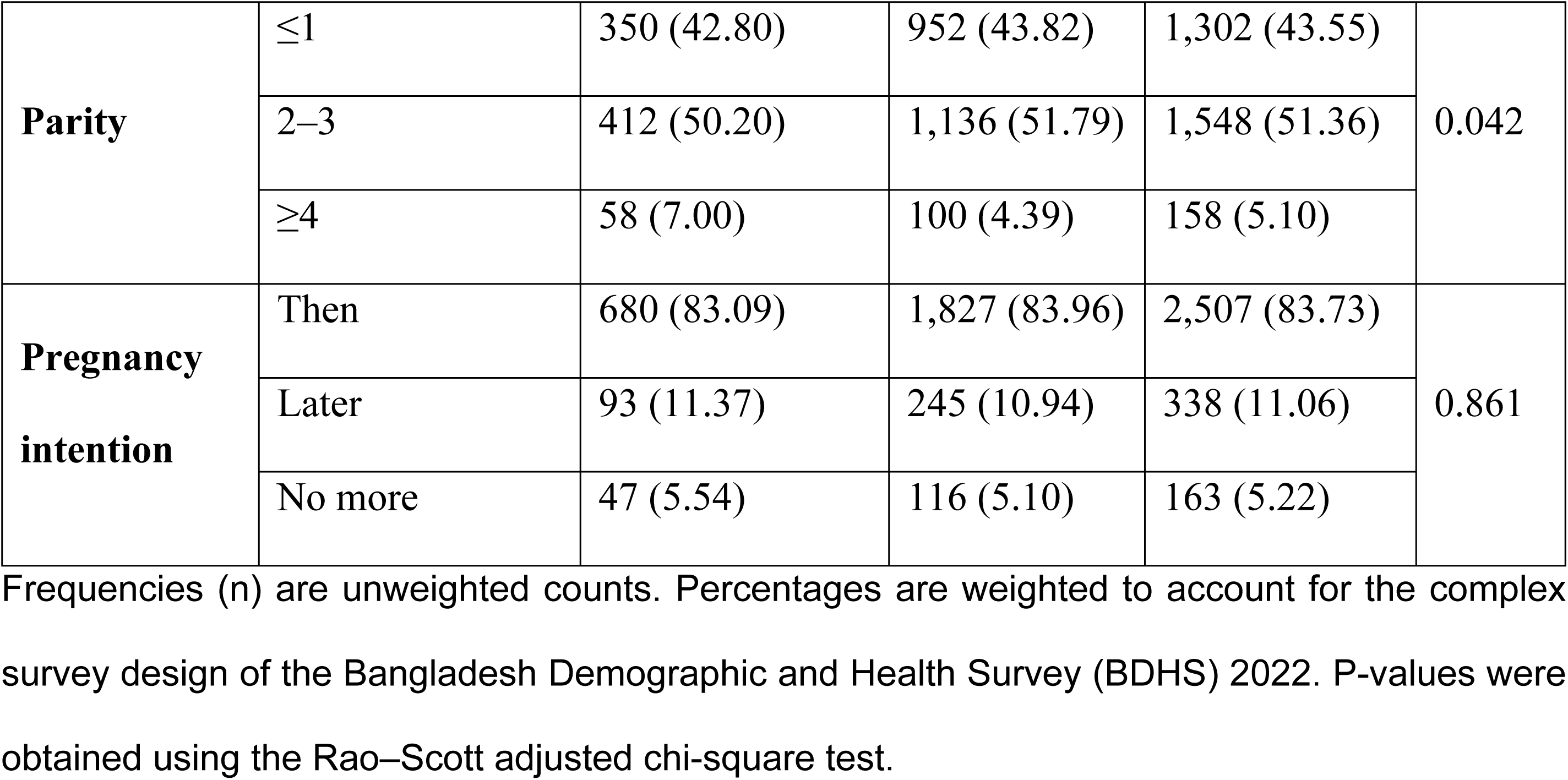
Characteristics of facility-based births by type of health facility, BDHS 2022.

Fig 2 shows the prevalence of cesarean delivery among facility-based births in Bangladesh. Overall, 71.2% of facility-based births were delivered by CS. The prevalence was substantially higher in private facilities (84.2%) than in public facilities (35.9%).

**Fig 2.**
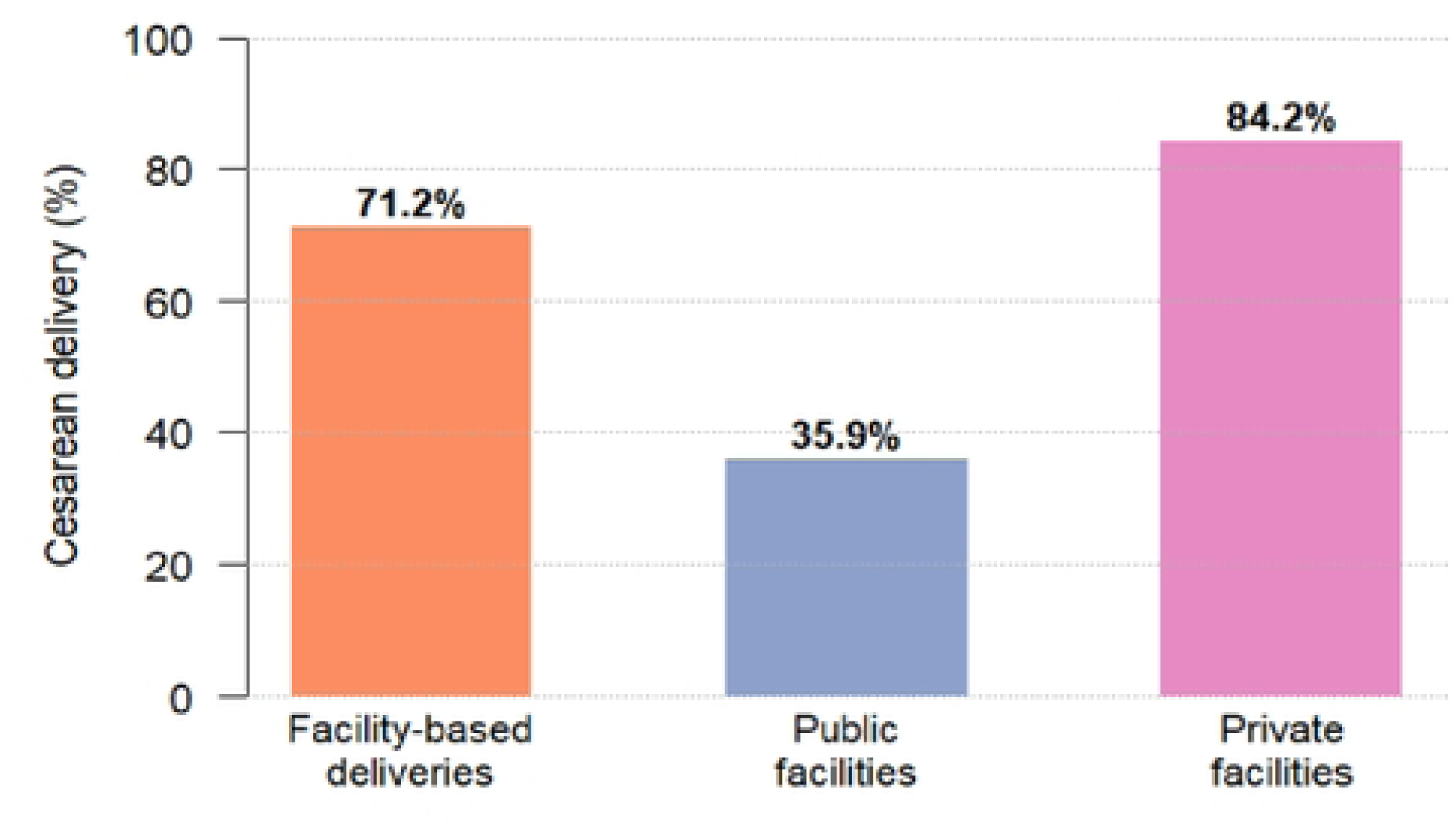
Percentage of cesarean deliveries among facility-based births in Bangladesh, by facility type.

### Factors associated with cesarean delivery among facility-based births

Table 2 presents the results of the survey-weighted multivariable logistic regression analysis. Compared with women delivering in public facilities, those delivering in private facilities had significantly higher odds of cesarean delivery (AOR: 9.07; 95% CI: 7.17–11.47). Wealth-related disparities were evident, with women from poorer (AOR: 1.53; 95% CI: 1.08–2.16), middle (AOR: 1.51; 95% CI: 1.07–2.12), and richest households (AOR: 1.92; 95% CI: 1.28–2.87) having higher odds of cesarean delivery than those from the poorest households. In contrast, women who reported that the pregnancy was unwanted had lower odds of cesarean delivery than those who wanted the pregnancy at that time (AOR: 0.56; 95% CI: 0.37–0.85). Significant regional differences were observed, with women residing in Chattogram (AOR: 0.43; 95% CI: 0.26–0.72) and Sylhet (AOR: 0.45; 95% CI: 0.26–0.76) divisions having lower odds of cesarean delivery compared with those residing in Barishal division. No significant associations were observed for maternal age, maternal education, place of residence, religion, husband’s education, husband’s occupation, media exposure, employment status, decision-making autonomy, distance to health facilities, ANC utilization, or parity.

**Table 2.** Socioeconomic, demographic, and reproductive factors associated with cesarean delivery among facility-based births in Bangladesh, BDHS 2022.

| Characteristics | AOR (95% CI) | p-value |
| --- | --- | --- |
| **Type of health facility** |  |  |
| Public | Ref | – |
| Private | 9.07 (7.17–11.47) | <0.001 |
| **Wealth quintile** |  |  |
| Poorest | Ref | – |
| Poorer | 1.53 (1.08–2.16) | 0.017 |
| Middle | 1.51 (1.07–2.12) | 0.019 |
| Richer | 1.32 (0.91–1.89) | 0.139 |
| Richest | 1.92 (1.28–2.87) | 0.002 |
| **Maternal age (years)** |  |  |
| 15–20 | Ref | – |
| 21–25 | 1.01 (0.74–1.39) | 0.929 |
| 26–30 | 1.13 (0.77–1.66) | 0.545 |
| ≥31 | 1.40 (0.86–2.28) | 0.176 |
| **Maternal education** |  |  |
| No education | Ref | – |
| Primary | 1.09 (0.62–1.94) | 0.758 |
| Secondary | 1.21 (0.69–2.13) | 0.512 |
| Higher | 1.33 (0.71–2.46) | 0.372 |
| **Place of residence** |  |  |
| Urban | Ref | – |
| Rural | 0.90 (0.68–1.18) | 0.441 |
| **Religion** |  |  |
| Islam | Ref | – |
| Other religions | 1.09 (0.74–1.60) | 0.656 |
| **Husband's education** |  |  |
| Less than secondary | Ref | – |
| Secondary and higher | 0.93 (0.73–1.18) | 0.547 |
| **Husband's occupation** |  |  |
| Not working | Ref | – |
| Agricultural worker | 0.79 (0.41–1.52) | 0.480 |
| Manual labor | 0.92 (0.50–1.70) | 0.786 |
| Professional/Service/Business | 0.95 (0.52–1.74) | 0.863 |
| **Exposure to mass media** |  |  |
| No | Ref | – |
| Yes | 1.01 (0.80–1.28) | 0.938 |
| **Currently employed** |  |  |
| No | Ref | – |
| Yes | 0.84 (0.64–1.11) | 0.222 |
| **Participation in household decision-making** |  |  |
| No | Ref | – |
| Yes | 1.30 (0.97–1.74) | 0.074 |
| **Distance to health facility is a problem** |  |  |
| No | Ref | – |
| Yes | 0.93 (0.75–1.16) | 0.532 |
| **ANC visits** |  |  |
| <4 visits | Ref | – |
| ≥4 visits | 1.01 (0.81–1.25) | 0.952 |
| **Parity** |  |  |
| ≤1 | Ref | – |
| 2–3 | 0.97 (0.74–1.28) | 0.849 |
| ≥4 | 0.67 (0.39–1.15) | 0.143 |
| **Pregnancy intention** |  |  |
| Wanted then | Ref | – |
| Wanted later | 0.82 (0.60–1.12) | 0.209 |
| Wanted no more | 0.56 (0.37–0.85) | 0.007 |
| **Division** |  |  |
| Barishal | Ref | – |
| Chattogram | 0.43 (0.26–0.72) | 0.001 |
| Dhaka | 1.19 (0.73–1.93) | 0.485 |
| Khulna | 1.48 (0.92–2.38) | 0.103 |
| Mymensingh | 1.42 (0.87–2.34) | 0.164 |
| Rajshahi | 1.07 (0.63–1.83) | 0.792 |
| Rangpur | 0.96 (0.61–1.52) | 0.866 |
| Sylhet | 0.45 (0.26–0.76) | 0.003 |

AOR = adjusted odds ratio; CI = confidence interval; Ref = reference category.

The determinants of cesarean delivery differed between public and private facilities (Table 3). In public facilities, women in the richest wealth quintile had significantly higher odds of cesarean delivery than those in the poorest quintile (AOR: 2.52; 95% CI: 1.35–4.73). Participation in household decision-making was also positively associated with cesarean delivery (AOR: 2.02; 95% CI: 1.23–3.30), while women reporting that the pregnancy was unwanted had lower odds of cesarean delivery (AOR: 0.44; 95% CI: 0.20–0.96). Significant regional variation was observed, with women residing in Chattogram (AOR: 0.40; 95% CI: 0.19–0.83), Rangpur (AOR: 0.30; 95% CI: 0.13–0.68), and Sylhet (AOR: 0.31; 95% CI: 0.13–0.72) divisions having lower odds of cesarean delivery than those residing in Barishal division.

**Table 3.** Factors associated with cesarean delivery among facility-based births by type of health facility, BDHS 2022.

| Characteristics | Public facilities |  | Private facilities |  |
| --- | --- | --- | --- | --- |
|  | AOR (95% CI) | p-value | AOR (95% CI) | p-value |
| **Wealth quintile** |  |  |  |  |
| Poorest | Ref | – | Ref | – |
| Poorer | 1.50 (0.86–2.62) | 0.154 | 1.72 (1.05–2.79) | 0.030 |
| Middle | 1.59 (0.88–2.86) | 0.123 | 1.52 (0.94–2.45) | 0.086 |
| Richer | 1.43 (0.81–2.54) | 0.217 | 1.33 (0.82–2.18) | 0.248 |
| Richest | 2.52 (1.35–4.73) | 0.004 | 1.67 (0.98–2.85) | 0.062 |
| **Maternal age (years)** |  |  |  |  |
| 15–20 | Ref | – | Ref | – |
| 21–25 | 0.91 (0.54–1.55) | 0.739 | 0.99 (0.66–1.49) | 0.966 |
| 26–30 | 1.04 (0.56–1.93) | 0.894 | 1.05 (0.62–1.77) | 0.856 |
| ≥31 | 1.04 (0.48–2.27) | 0.916 | 1.53 (0.82–2.83) | 0.179 |
| **Maternal education** |  |  |  |  |
| No education | Ref | – | Ref | – |
| Primary | 1.04 (0.44–2.42) | 0.936 | 1.26 (0.61–2.59) | 0.533 |
| Secondary | 1.21 (0.54–2.72) | 0.647 | 1.37 (0.68–2.79) | 0.378 |
| Higher | 1.15 (0.45–2.95) | 0.769 | 1.64 (0.74–3.67) | 0.223 |
| **Place of residence** |  |  |  |  |
| Urban | Ref | – | Ref | – |
| Rural | 0.72 (0.48–1.08) | 0.107 | 1.01 (0.71–1.45) | 0.943 |
| **Religion** |  |  |  |  |
| Islam | Ref | – | Ref | – |
| Other religions | 1.21 (0.68–2.16) | 0.511 | 1.03 (0.58–1.81) | 0.931 |
| **Husband's education** |  |  |  |  |
| Less than secondary | Ref | – | Ref | – |
| Secondary and higher | 0.80 (0.54–1.18) | 0.262 | 1.04 (0.74–1.46) | 0.842 |
| **Husband's occupation** |  |  |  |  |
| Not working | Ref | – | Ref | – |
| Agricultural worker | 3.02 (0.81–11.22) | 0.098 | 0.37 (0.12–1.09) | 0.072 |
| Manual labor | 1.71 (0.48–6.10) | 0.407 | 0.67 (0.23–1.94) | 0.460 |
| Professional/Service/Business | 3.02 (0.86–10.66) | 0.085 | 0.46 (0.16–1.28) | 0.136 |
| **Exposure to mass media** |  |  |  |  |
| No | Ref | – | Ref | – |
| Yes | 0.87 (0.57–1.33) | 0.521 | 1.09 (0.83–1.44) | 0.543 |
| **Currently employed** |  |  |  |  |
| No | Ref | – | Ref | – |
| Yes | 1.03 (0.64–1.64) | 0.914 | 0.71 (0.50–1.01) | 0.059 |
| **Participation in household decision-making** |  |  |  |  |
| No | Ref | – | Ref | – |
| Yes | 2.02 (1.23–3.30) | 0.005 | 1.02 (0.67–1.56) | 0.920 |
| **Distance to health facility is a problem** |  |  |  |  |
| No | Ref | – | Ref | – |
| Yes | 1.06 (0.73–1.52) | 0.772 | 0.84 (0.64–1.09) | 0.192 |
| **ANC visits** |  |  |  |  |
| <4 visits | Ref | – | Ref | – |
| ≥4 visits | 0.87 (0.59–1.28) | 0.473 | 1.13 (0.87–1.47) | 0.367 |
| **Parity** |  |  |  |  |
| ≤1 | Ref | – | Ref | – |
| 2–3 | 0.94 (0.59–1.51) | 0.811 | 1.07 (0.75–1.54) | 0.708 |
| ≥4 | 0.75 (0.31–1.82) | 0.529 | 0.57 (0.30–1.11) | 0.097 |
| **Pregnancy intention** |  |  |  |  |
| Wanted then | Ref | – | Ref | – |
| Wanted later | 0.74 (0.43–1.27) | 0.279 | 0.84 (0.54–1.29) | 0.421 |
| Wanted no more | 0.44 (0.20–0.96) | 0.039 | 0.63 (0.37–1.07) | 0.084 |
| **Division** |  |  |  |  |
| Barishal | Ref | – | Ref | – |
| Chattogram | 0.40 (0.19–0.83) | 0.014 | 0.46 (0.25–0.84) | 0.012 |
| Dhaka | 0.79 (0.37–1.68) | 0.544 | 1.34 (0.76–2.35) | 0.309 |
| Khulna | 0.56 (0.25–1.24) | 0.154 | 2.71 (1.48–4.94) | 0.001 |
| Mymensingh | 0.46 (0.21–1.02) | 0.057 | 4.31 (2.09–8.88) | <0.001 |
| Rajshahi | 0.77 (0.32–1.84) | 0.560 | 1.29 (0.71–2.32) | 0.406 |
| Rangpur | 0.30 (0.13–0.68) | 0.004 | 1.76 (0.96–3.23) | 0.069 |
| Sylhet | 0.31 (0.13–0.72) | 0.007 | 0.52 (0.28–0.96) | 0.037 |

AOR = adjusted odds ratio; CI = confidence interval; Ref = reference category.

In private facilities, wealth remained an important determinant of cesarean delivery. Women from poorer households had significantly higher odds of cesarean delivery than those from the poorest households (AOR: 1.72; 95% CI: 1.05–2.79). Although women from the middle and richest wealth quintiles also exhibited elevated odds of cesarean delivery, these associations did not reach statistical significance. Maternal education was not significantly associated with cesarean delivery in private facilities. Significant regional variation was observed, with lower odds of cesarean delivery among women residing in Chattogram (AOR: 0.46; 95% CI: 0.25–0.84) and Sylhet (AOR: 0.52; 95% CI: 0.28–0.96) divisions and higher odds among those residing in Khulna (AOR: 2.71; 95% CI: 1.48–4.94) and Mymensingh (AOR: 4.31; 95% CI: 2.09–8.88) divisions compared with Barishal division. No significant associations were observed for maternal age, maternal education, residence, religion, husband’s education, husband’s occupation, media exposure, employment status, decision-making autonomy, distance to health facilities, ANC utilization, parity, or pregnancy intention.

### Inequality in C-section delivery

Fig 3 illustrates the prevalence of cesarean delivery across household wealth quintiles by facility type. Overall, cesarean delivery prevalence increased progressively with household wealth, rising from 57.8% among women in the poorest wealth quintile to 79.7% among those in the richest quintile. A similar socioeconomic gradient was observed in both public and private facilities. The increase was particularly pronounced among public facility deliveries, where the prevalence increased from 23.6% in the poorest wealth quintile to 51.6% in the richest wealth quintile. In private facilities, cesarean delivery prevalence remained consistently high across all wealth groups, ranging from 78.4% among the poorest women to 87.2% among the richest women.

**Fig 3.**
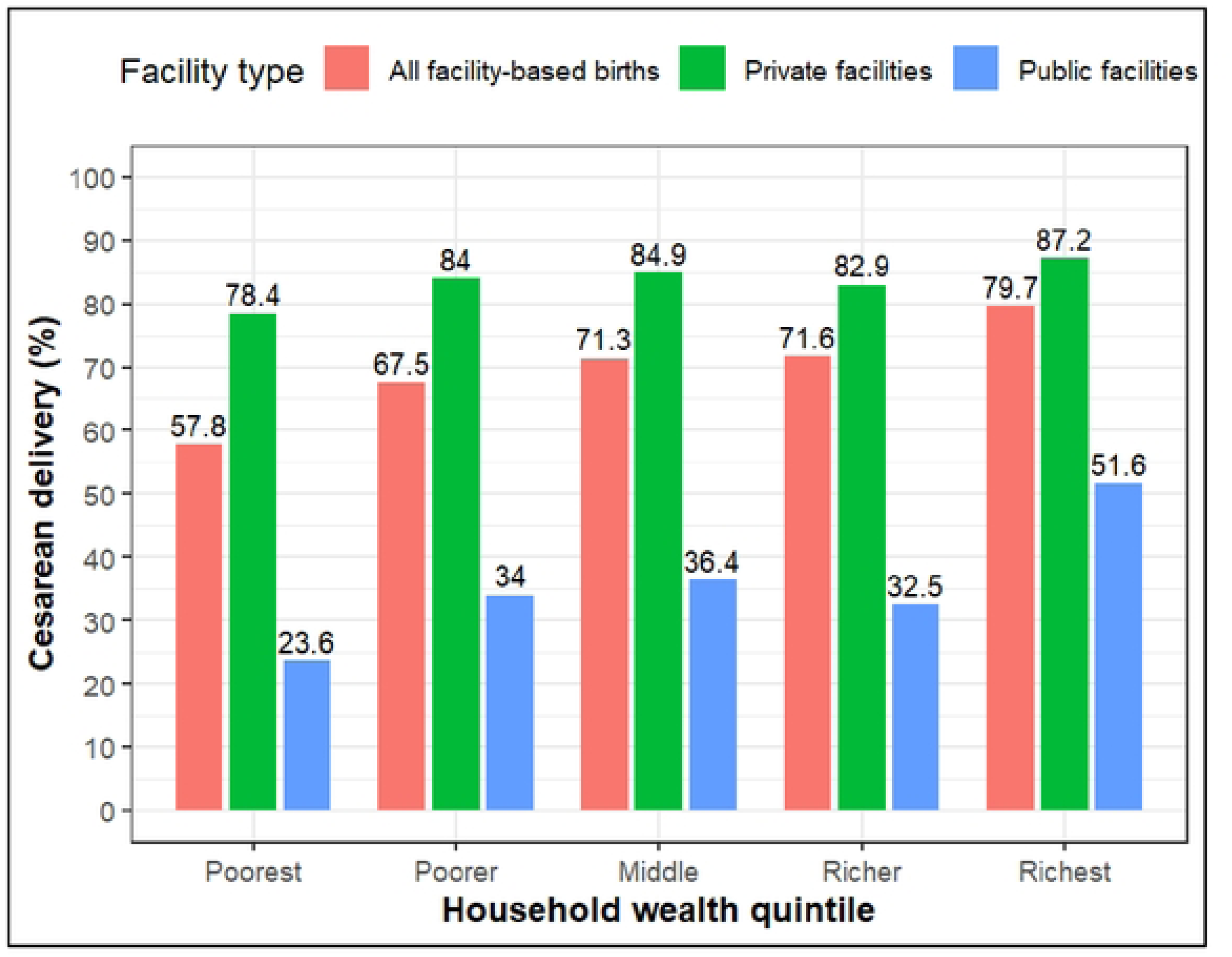
Prevalence of cesarean delivery across household wealth quintiles by facility type in Bangladesh, BDHS 2022.

Facility-specific prevalence estimates across socioeconomic, demographic, and reproductive characteristics are presented in Supplementary Table S1. In public facilities, cesarean delivery prevalence varied substantially across wealth quintiles, place of residence, divisions, husband’s occupation, decision-making autonomy, and pregnancy intention, whereas prevalence remained consistently high across most subgroups in private facilities.

Fig 4 presents the concentration curves for cesarean delivery by facility type. All concentration curves lay below the line of equality, indicating that cesarean delivery was disproportionately concentrated among women from wealthier households. The greatest deviation from the line of equality was observed for public facilities, whereas the private facility curve was closest to the equality line, with the overall curve positioned between the two. Consistent with these patterns, the Erreygers-corrected concentration index (ECCI) was 0.154 (95% CI: 0.117–0.191) for all facility-based births (Table 4). The magnitude of inequality was substantially higher among women delivering in public facilities (ECCI = 0.189; 95% CI: 0.114–0.264) than among those delivering in private facilities (ECCI = 0.049; 95% CI: 0.014–0.084), corresponding to an absolute difference of 0.140. All ECCI estimates were positive and statistically significant, indicating that cesarean delivery was disproportionately concentrated among women from wealthier households across all facility types. The substantially higher ECCI observed in public facilities suggests a stronger socioeconomic gradient in cesarean delivery utilization within the public sector than within the private sector.

**Fig 4.**
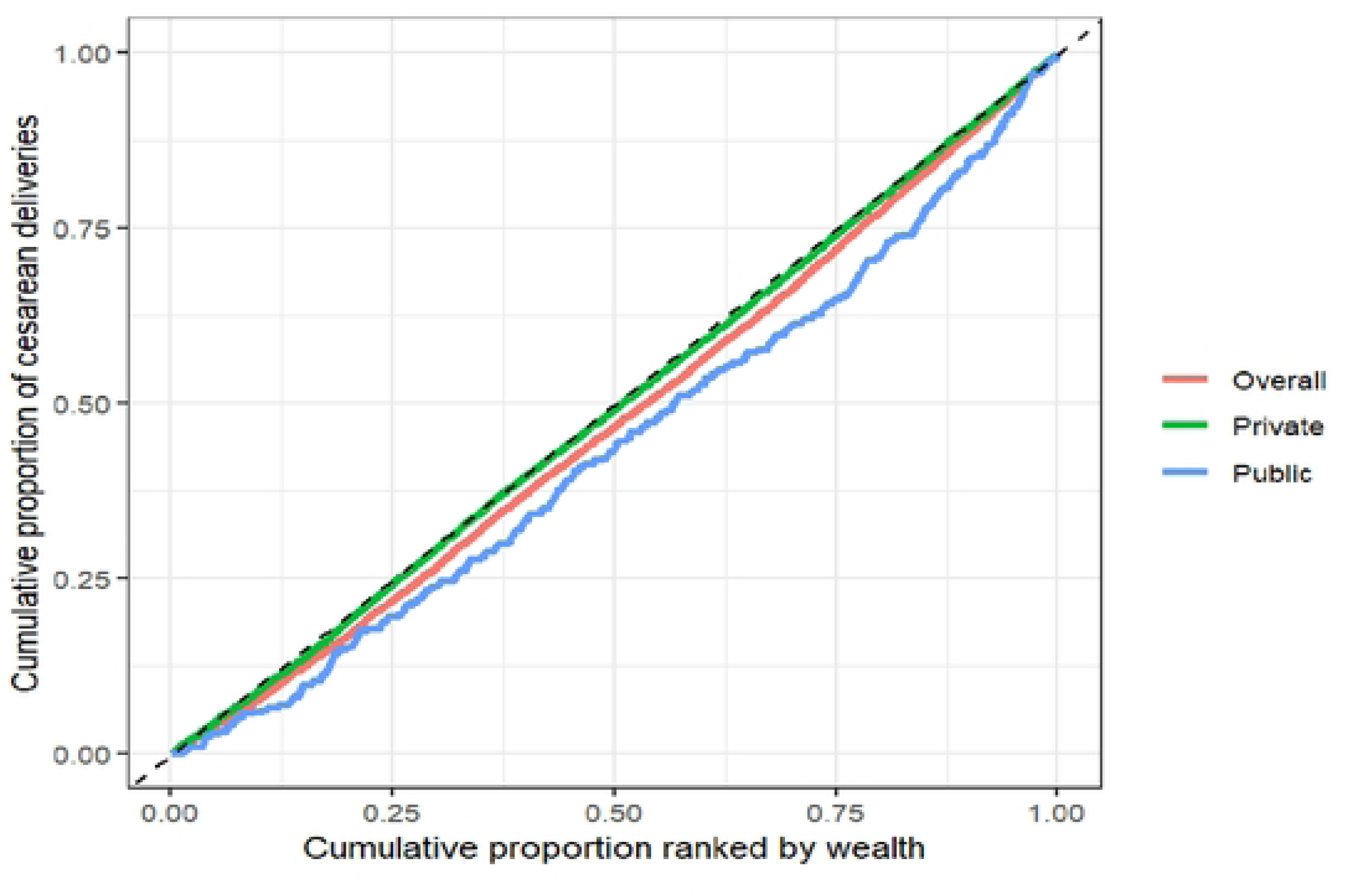
Concentration curves of cesarean delivery among facility-based births by facility type in Bangladesh, BDHS 2022.

**Table 4.** Wealth-related inequality in cesarean delivery among facility-based births by type of health facility, BDHS 2022.

| Facility type | ECCI | 95% CI | p-value |
| --- | --- | --- | --- |
| Overall | 0.154 | 0.117 to 0.191 | <0.001 |
| Public facilities | 0.189 | 0.114 to 0.264 | <0.001 |
| Private facilities | 0.049 | 0.014 to 0.084 | 0.007 |
| Difference (Public – Private) | 0.140 | – | – |

ECCI = Erreygers-corrected concentration index for the binary outcome (cesarean delivery). Positive values indicate that cesarean delivery is disproportionately concentrated among women from wealthier households (pro-rich inequality), whereas negative values indicate concentration among poorer households (pro-poor inequality).

### Decomposition analysis

Decomposition was performed using the standard concentration index, whereas inequality was summarized using the Erreygers-corrected concentration index because the outcome was binary.

The decomposition analysis indicated substantial pro-rich socioeconomic inequality in cesarean delivery among all facility-based births (CI = 0.054; 95% CI: 0.041–0.067), with considerably greater inequality observed in public facilities (CI = 0.133; 95% CI: 0.080–0.185) than in private facilities (CI = 0.015; 95% CI: 0.004–0.025). The included determinants explained 97.5%, 97.6%, and 91.5% of the observed inequality among all facility-based births, public facilities, and private facilities, respectively, with residual contributions of 2.5%, 2.4%, and 8.5% (Table 5).

**Table 5.** Decomposition of wealth-related socioeconomic inequality in cesarean delivery among facility-based births by type of health facility, BDHS 2022.

| Characteristics | Overall |  |  |  | Public |  |  |  | Private |  |  |  |
| --- | --- | --- | --- | --- | --- | --- | --- | --- | --- | --- | --- | --- |
|  | E | CIx | C | PC (%) | E | CIx | C | PC (%) | E | CIx | C | PC (%) |
| **Maternal age (years)** |  |  |  |  |  |  |  |  |  |  |  |  |
| 21–25 | -0.0054 | -0.0226 | 0.0001 | 0.22 | ̄<br>0.0151 | ̄<br>0.0226 | 0.0003 | 0.26 | ̄<br>0.0003 | ̄<br>0.0136 | 0.0000 | 0.03 |
| 26–30 | 0.0091 | 0.0471 | 0.0004 | 0.78 | 0.0062 | 0.0520 | 0.0003 | 0.24 | 0.0022 | 0.0415 | 0.0001 | 0.63 |
| ≥31 | 0.0139 | 0.1117 | 0.0016 | 2.86 | 0.0062 | 0.0432 | 0.0003 | 0.20 | 0.0110 | 0.1320 | 0.0014 | 9.96 |
| **Maternal education** |  |  |  |  |  |  |  |  |  |  |  |  |
| Primary | 0.0120 | -0.2739 | ̄<br>0.0033 | -6.05 | 0.0067 | ̄<br>0.2336 | ̄<br>0.0016 | -1.17 | 0.0058 | ̄<br>0.2835 | ̄<br>0.0016 | -11.34 |
| Secondary | 0.0597 | -0.0265 | ̄<br>0.0016 | -2.91 | 0.0657 | 0.0141 | 0.0009 | 0.70 | 0.0302 | ̄<br>0.0405 | ̄<br>0.0012 | -8.43 |
| Higher | 0.0331 | 0.3011 | 0.0100 | 18.30 | 0.0160 | 0.3168 | 0.0051 | 3.82 | 0.0202 | 0.2851 | 0.0058 | 39.64 |
| **Place of residence** |  |  |  |  |  |  |  |  |  |  |  |  |
| Rural | 0.0049 | -0.1471 | ̄<br>0.0007 | -1.33 | ̄<br>0.1491 | ̄<br>0.1505 | 0.0224 | 16.91 | 0.0046 | ̄<br>0.1454 | ̄<br>0.0007 | -4.59 |
| **Religion** |  |  |  |  |  |  |  |  |  |  |  |  |
| Other religions | 0.0032 | -0.0884 | ̄<br>0.0003 | -0.52 | 0.0103 | ̄<br>0.0561 | ̄<br>0.0006 | -0.44 | ̄<br>0.0006 | ̄<br>0.1007 | 0.0001 | 0.40 |
| **Husband's education** |  |  |  |  |  |  |  |  |  |  |  |  |
| Secondary and higher | -0.0004 | 0.1482 | ̄<br>0.0001 | -0.12 | ̄<br>0.0734 | 0.1516 | ̄<br>0.0111 | -8.40 | 0.0060 | 0.1405 | 0.0008 | 5.79 |
| **Husband's occupation** |  |  |  |  |  |  |  |  |  |  |  |  |
| Agricultural worker | -0.0038 | -0.2907 | 0.0011 | 2.03 | 0.0880 | ̄<br>0.2746 | ̄<br>0.0242 | -18.22 | ̄<br>0.0198 | ̄<br>0.2975 | 0.0059 | 40.40 |
| Manual labor | -0.0103 | -0.1562 | 0.0016 | 2.95 | 0.0978 | ̄<br>0.1024 | ̄<br>0.0100 | -7.55 | ̄<br>0.0099 | ̄<br>0.1648 | 0.0016 | 11.25 |
| Professional/Service/Business | 0.0110 | 0.1506 | 0.0017 | 3.05 | 0.2744 | 0.1520 | 0.0417 | 31.45 | ̄<br>0.0543 | 0.1409 | ̄<br>0.0077 | -52.63 |
| **Media exposure** |  |  |  |  |  |  |  |  |  |  |  |  |
| Yes | -0.0073 | 0.1217 | ̄<br>0.0009 | -1.64 | ̄<br>0.0458 | 0.1394 | ̄<br>0.0064 | -4.81 | 0.0098 | 0.1132 | 0.0011 | 7.66 |
| **Currently employed** |  |  |  |  |  |  |  |  |  |  |  |  |
| Yes | -0.0120 | -0.1083 | 0.0013 | 2.39 | 0.0024 | ̄<br>0.0938 | ̄<br>0.0002 | -0.17 | ̄<br>0.0092 | ̄<br>0.1118 | 0.0010 | 7.03 |
| **Wealth quintile** |  |  |  |  |  |  |  |  |  |  |  |  |
| Poorer | 0.0257 | -0.5575 | <sup>-</sup>0.0143 | -26.33 | 0.0458 | <sup>-</sup>0.4073 | <sup>-</sup>0.0187 | -14.07 | 0.0147 | <sup>-</sup>0.6146 | <sup>-</sup>0.0090 | -62.16 |
| Middle | 0.0331 | -0.1679 | <sup>-</sup>0.0056 | -10.23 | 0.0523 | 0.0198 | 0.0010 | 0.78 | 0.0147 | <sup>-</sup>0.2393 | <sup>-</sup>0.0035 | -24.25 |
| Richer | 0.0389 | 0.2743 | 0.0107 | 19.59 | 0.0363 | 0.4249 | 0.0154 | 11.63 | 0.0126 | 0.2171 | 0.0027 | 18.87 |
| Richest | 0.0588 | 0.7544 | 0.0443 | 81.49 | 0.1045 | 0.8096 | 0.0846 | 63.77 | 0.0218 | 0.7335 | 0.0160 | 109.69 |
| **Decision-making autonomy** |  |  |  |  |  |  |  |  |  |  |  |  |
| Yes | 0.0796 | 0.0059 | 0.0005 | 0.86 | 0.3226 | 0.0033 | 0.0011 | 0.79 | 0.0058 | 0.0055 | 0.0000 | 0.22 |
| **Distance to health facility is a problem** |  |  |  |  |  |  |  |  |  |  |  |  |
| Yes | -0.0049 | -0.0454 | 0.0002 | 0.41 | 0.0125 | <sup>-</sup>0.0646 | <sup>-</sup>0.0008 | -0.61 | <sup>-</sup>0.0113 | <sup>-</sup>0.0364 | 0.0004 | 2.82 |
| **ANC visits** |  |  |  |  |  |  |  |  |  |  |  |  |
| ≥4 visits | 0.0103 | 0.1544 | 0.0016 | 2.92 | <sup>-</sup>0.0382 | 0.1684 | <sup>-</sup>0.0064 | -4.86 | 0.0084 | 0.1448 | 0.0012 | 8.34 |
| **Parity** |  |  |  |  |  |  |  |  |  |  |  |  |
| 2–3 | -0.0059 | 0.0250 | <sup>-</sup>0.0001 | -0.27 | <sup>-</sup>0.0158 | 0.0079 | <sup>-</sup>0.0001 | -0.09 | 0.0047 | 0.0317 | 0.0002 | 1.04 |
| ≥4 | -0.0083 | -0.1502 | 0.0012 | 2.29 | <sup>-</sup>0.0140 | <sup>-</sup>0.1473 | 0.0021 | 1.56 | <sup>-</sup>0.0051 | <sup>-</sup>0.1359 | 0.0007 | 4.73 |
| **Pregnancy intention** |  |  |  |  |  |  |  |  |  |  |  |  |
| Wanted later | -0.0051 | -0.0551 | 0.0003 | 0.52 | <sup>-</sup>0.0202 | 0.0098 | <sup>-</sup>0.0002 | -0.15 | <sup>-</sup>0.0028 | <sup>-</sup>0.0771 | 0.0002 | 1.46 |
| Wanted no more | -0.0062 | -0.0689 | 0.0004 | 0.79 | <sup>-</sup>0.0221 | <sup>-</sup>0.1670 | 0.0037 | 2.79 | <sup>-</sup>0.0036 | <sup>-</sup>0.0241 | 0.0001 | 0.59 |
| **Division** |  |  |  |  |  |  |  |  |  |  |  |  |
| Chattogram | -0.0559 | -0.0180 | 0.0010 | 1.85 | <sup>-</sup>0.1397 | <sup>-</sup>0.0144 | 0.0020 | 1.52 | <sup>-</sup>0.0279 | 0.0031 | <sup>-</sup>0.0001 | -0.60 |
| Dhaka | 0.0060 | 0.2249 | 0.0013 | 2.47 | <sup>-</sup>0.0283 | 0.2753 | <sup>-</sup>0.0078 | -5.87 | 0.0124 | 0.1990 | 0.0025 | 16.98 |
| Khulna | 0.0111 | 0.0699 | 0.0008 | 1.42 | <sup>-</sup>0.0381 | 0.1015 | <sup>-</sup>0.0039 | -2.92 | 0.0180 | 0.0546 | 0.0010 | 6.76 |
| Mymensingh | 0.0029 | -0.2173 | <sup>-</sup>0.0006 | -1.18 | <sup>-</sup>0.0399 | <sup>-</sup>0.2563 | 0.0102 | 7.70 | 0.0114 | <sup>-</sup>0.1899 | <sup>-</sup>0.0022 | -14.92 |
| Rajshahi | 0.0069 | -0.0486 | <sup>-</sup>0.0003 | -0.62 | <sup>-</sup>0.0104 | 0.0754 | <sup>-</sup>0.0008 | -0.59 | 0.0050 | <sup>-</sup>0.0932 | <sup>-</sup>0.0005 | -3.21 |
| Rangpur | -0.0021 | -0.2840 | 0.0006 | 1.07 | <sup>-</sup>0.0743 | <sup>-</sup>0.3179 | 0.0236 | 17.81 | 0.0091 | <sup>-</sup>0.2692 | <sup>-</sup>0.0025 | -16.87 |
| Sylhet | -0.0153 | -0.0157 | 0.0002 | 0.44 | -0.0492 | -0.1510 | 0.0074 | 5.60 | -0.0054 | 0.1016 | -0.0006 | -3.80 |
| **Residual** | — | — | 0.0014 | 2.48 | — | — | 0.0031 | 2.37 | — | — | 0.0012 | 8.50 |
| **Total** | — | — | **0.0544** | **100.00** | — | — | **0.1326** | **100.00** | — | — | **0.0145** | **100.00** |

E = elasticity; Clx = concentration index of determinant; C = absolute contribution to the concentration index of cesarean delivery; PC = percentage contribution. Positive percentage contributions indicate factors increasing pro-rich inequality, whereas negative contributions indicate factors reducing pro-rich inequality.

Among all facility-based births, household wealth was the dominant contributor to socioeconomic inequality in cesarean delivery. The richest wealth quintile alone accounted for 81.5% of the observed inequality, followed by the richer wealth quintile (19.6%) and higher maternal education (18.3%). Additional positive contributions were observed for husband’s occupation (Professional/Service/Business: 3.1%; manual labor: 3.0%; agricultural worker: 2.0%), antenatal care utilization (2.9%), maternal age ≥31 years (2.9%), employment (2.4%), and parity of four or more children (2.3%). In contrast, the poorer (−26.3%) and middle (−10.2%) wealth quintiles and primary maternal education (−6.1%) reduced the magnitude of the observed pro-rich inequality (Table 5).

In public facilities, the largest contributors to inequality were the richest wealth quintile (63.8%), husbands’ Professional/Service/Business occupation (31.5%), rural residence (16.9%), Rangpur division (17.8%), and richer wealth status (11.6%). Additional positive contributions were observed for Mymensingh (7.7%) and Sylhet (5.6%) divisions. Factors reducing inequality included agricultural occupation of husbands (−18.2%), poorer wealth status (−14.1%), husband’s secondary or higher education (−8.4%), manual labor occupation of husbands (−7.6%), residence in Dhaka division (−5.9%), and having four or more ANC visits (−4.9%) (Table 5).

In private facilities, the richest wealth quintile remained the largest contributor, accounting for 109.7% of the observed inequality. Other major contributors included agricultural occupation of husbands (40.4%), higher maternal education (39.6%), richer wealth status (18.9%), Dhaka division (17.0%), manual labor occupation of husbands (11.3%), maternal age ≥31 years (10.0%), antenatal care utilization (8.3%), media exposure (7.7%), employment (7.0%), Khulna division (6.8%), and husband’s secondary or higher education (5.8%). Conversely, Professional/Service/Business occupation of husbands (−52.6%), poorer (−62.2%) and middle (−24.3%) wealth quintiles, Rangpur (−16.9%) and Mymensingh (−14.9%) divisions, primary maternal education (−11.3%), secondary maternal education (−8.4%), and rural residence (−4.6%) reduced the magnitude of the observed pro-rich inequality (Table 5).

## Discussion

Using nationally representative data from the BDHS 2022, this study examined wealth-related inequalities in CS utilization among facility-based births in Bangladesh and explored how these inequalities differ across public and private healthcare facilities. Three major findings emerged. First, CS utilization among facility births was exceptionally high, particularly in private facilities. Second, substantial pro-rich socioeconomic inequalities in CS utilization persisted, with CS disproportionately concentrated among women from wealthier households. Third, and most importantly, the magnitude and drivers of inequality differed markedly between public and private facilities, highlighting the importance of considering health-system context when assessing inequities in obstetric care.

The overall prevalence of CS among facility-based births exceeded 70%, while more than four-fifths of births in private facilities were delivered by cesarean section. These findings reinforce concerns regarding the continuing escalation of CS use in Bangladesh, a country that has experienced one of the fastest increases in CS utilization globally [13]. Although increased availability of obstetric services may have improved access to life-saving interventions for women experiencing delivery complications, the extremely high prevalence observed in private facilities raises concerns about the potential overuse of CS [13]. Previous studies from Bangladesh have suggested that financial incentives, provider preferences, institutional norms, and increasing demand for scheduled deliveries may contribute to elevated CS rates in private-sector facilities [14, 15]. Consequently, the challenge facing Bangladesh is no longer solely one of access but also one of ensuring the appropriate use of CS based on clinical need [16].

Consistent with previous evidence from Bangladesh and other low– and middle-income countries, we found that CS utilization was disproportionately concentrated among women from wealthier households. Similar pro-rich inequalities have been reported using concentration indices, concentration curves, and other equity measures in Bangladesh, Ghana, and numerous LMIC settings [4–6]. The persistence of these disparities suggests that economic advantage continues to play a critical role in shaping access to obstetric services. Wealthier women may be better positioned to afford direct and indirect healthcare costs, access higher-quality facilities, seek specialist care, and exercise greater autonomy in healthcare decision-making. At the same time, poorer women may continue to face financial, informational, and geographic barriers that limit access to medically necessary cesarean delivery. Therefore, rising national CS rates should not be interpreted as evidence of equitable access to obstetric care.

The nearly fourfold higher concentration index observed in public facilities compared with private facilities suggests that socioeconomic differences in access to cesarean delivery remain substantially more pronounced within the public sector. At first glance, this finding appears counterintuitive because private healthcare services are generally more expensive and tend to serve more affluent populations. However, the lower concentration index observed in private facilities does not necessarily indicate greater equity. Rather, it reflects the fact that CS utilization was uniformly high across all wealth groups among women who delivered in private facilities. In contrast, CS utilization within public facilities exhibited a much stronger socioeconomic gradient, with considerably higher rates among wealthier women than among poorer women. This pattern suggests that inequalities in access to cesarean delivery remain deeply embedded within the public healthcare system, despite its mandate to provide affordable and equitable maternal healthcare services. Wealthier women may possess greater resources and knowledge to navigate referral pathways, obtain specialist consultations, and access higher-quality services within public facilities, whereas poorer women may continue to encounter barriers even when financial costs are relatively low [17].

The decomposition analysis further revealed that household wealth was the dominant contributor to observed inequalities in both public and private facilities. This finding is consistent with previous decomposition studies showing that economic status remains the principal driver of disparities in maternal healthcare utilization [18–20]. However, the pathways through which inequalities were generated differed across facility types. In public facilities, wealth-related disparities were reinforced by geographic and contextual factors, including rural residence, regional variation, and occupational characteristics, suggesting the continued importance of structural barriers in accessing cesarean delivery. In contrast, inequalities within private facilities were primarily driven by household wealth, with additional contributions from higher maternal education and occupational characteristics. Notably, although maternal education was not independently associated with cesarean delivery after adjustment for other factors, higher education contributed substantially to wealth-related inequality because educational attainment remained disproportionately concentrated among wealthier women. These findings indicate that wealth-related inequalities are produced through multiple pathways that differ across segments of the healthcare system.

The observed regional variations further highlight the importance of local health-system contexts. Significant differences across divisions suggest that the availability of obstetric services, referral practices, healthcare infrastructure, and provider behavior may vary substantially throughout the country. Similar geographical inequalities have been documented in previous studies of maternal healthcare utilization in Bangladesh and indicate that national averages may conceal important subnational disparities [21, 22]. Future research should therefore explore contextual and health-system factors that contribute to regional variation in cesarean delivery practices.

The findings have several policy implications. Efforts to address inequalities in CS utilization should pursue a dual objective: improving access to medically necessary cesarean delivery among disadvantaged populations while simultaneously reducing unnecessary procedures among socioeconomically advantaged groups. Within the public sector, interventions aimed at reducing geographic and socioeconomic barriers, strengthening referral systems, and improving equitable access to emergency obstetric care may help narrow wealth-related disparities in cesarean delivery. In the private sector, where cesarean delivery rates were uniformly high across wealth groups, greater emphasis should be placed on strengthening regulation, clinical auditing, and adherence to evidence-based indications for cesarean delivery. Monitoring CS utilization by both socioeconomic status and facility type may provide a more informative assessment of equity than national averages alone.

This study has several strengths. It utilized the most recent nationally representative data from Bangladesh and applied concentration index and decomposition techniques to quantify both the magnitude and sources of wealth-related inequality. Moreover, by conducting separate analyses for public and private facilities, the study provides novel evidence on facility-specific patterns of inequality that have received limited attention in previous research. However, several limitations should be acknowledged. The BDHS lacks information on clinical indications for cesarean delivery, preventing assessment of whether procedures were medically justified. Furthermore, provider-level and facility-level factors that may influence delivery decisions were not available in the survey. Despite these limitations, the study offers important insights into the socioeconomic distribution of cesarean delivery in Bangladesh and the mechanisms underlying observed inequalities.

In conclusion, wealth-related inequalities in cesarean delivery remain substantial in Bangladesh despite widespread increases in CS utilization. Although pro-rich inequality exists across both public and private healthcare sectors, the magnitude and underlying drivers of inequality differ considerably between facility types. These findings suggest that policies aimed at improving equity in maternal healthcare should move beyond national-level assessments and consider the distinct pathways through which inequalities are generated within different segments of the healthcare system.

## Data Availability

The data underlying the results presented in this study are publicly available from the Demographic and Health Surveys (DHS) Program. The 2022 Bangladesh Demographic and Health Survey (BDHS) dataset can be obtained upon reasonable request and approval from the DHS Program through the DHS website (https://dhsprogram.com/data/available-datasets.cfm). The authors did not receive special access privileges to these data.

---

Supplementary Table S1

